# First-in-human intrapulmonary intratarget microdosing of a novel dual inflammasome inhibitor of NLRP 1/ NLRP 3 in ex vivo human lungs and patients with interstitial lung disease

**DOI:** 10.64898/2026.05.05.26352329

**Authors:** Tom Quinn, Feng Li, Becky Wheeler, Stuart Dickson, Katie Hamilton, Anuruddika Fernando, Charles Lochenie, Joanne Mair, Sarah McNamara, Karen Linton, Erin Gaughan, Richard O’Connor, Antonella Pellicoro, Kay Russell, Annya Bruce, Scott Denham, Natalie Homer, Ashley Mansell, Manu Shankar-Hari, Adriano Rossi, Ahsan Akram, Keith Finlayson, Nik Hirani, Kevin Dhaliwal

## Abstract

The development of lung-directed therapeutics is limited by poor translational fidelity between preclinical models and early-phase clinical trials. We report a first-in-human Phase 0 intratarget microdosing study demonstrating the feasibility of intrapulmonary delivery and pharmacological interrogation of a novel inflammasome inhibitor. A 100 μg microdose of ADS032, a dual NLRP1/NLRP3 inhibitor, was administered to distal airways via bronchoscopy in patients with interstitial lung disease, informed by optimisation in *ex vivo* human lung perfusion and ventilation systems. Clinical-grade manufacture, formulation, stability, and toxicology enabled intrapulmonary administration. Using liquid chromatography–mass spectrometry, ADS032 was detected in plasma, bronchoalveolar lavage fluid, distal airway micro-aspirates, and recovered cells, with spatially resolved sampling achieved without cross-contamination. Fluorescent labelling enabled direct visualisation of alveolar drug uptake *ex vivo*. These findings establish intrapulmonary intratarget microdosing as a human-relevant platform for early pharmacological evaluation of lung therapeutics prior to Phase 1 trials.

## Introduction

The translation of anti-inflammatory therapeutics to the clinic remains a major challenge in respiratory medicine, driven in part by limited predictive value of preclinical models and late-stage clinical attrition[1, 2]. This challenge is particularly relevant for lung-directed therapies, where systemic administration often fails to achieve sufficient local exposure or results in dose-limiting off-target effects. As a result, promising mechanistic candidates are frequently abandoned before meaningful evaluation of target engagement in the human lung.

Phase 0 clinical trials offer a regulatory framework to address this translational gap by enabling early assessment of pharmacokinetics and pharmacodynamics using subtherapeutic drug doses in humans[3–5]. However, conventional Phase 0 designs typically rely on systemic administration and peripheral sampling, providing limited insight into drug behaviour within the target organ. In contrast, intratarget microdosing (ITM) enables controlled delivery of microdoses directly to a defined anatomical compartment, allowing local drug exposure, spatially resolved sampling, and early mechanistic readouts while remaining within stringent safety margins[6, 7].

Here, we present a first-in-human application of intrapulmonary intratarget microdosing to evaluate a novel inflammasome inhibitor termed ADS032[8]. The conduction of lung microdosing experiments with an inflammasome inhibitor is important to evaluate localized inflammatory activity in the respiratory system, where inflammasome-driven inflammation contributes to ARDS, COPD, and pulmonary fibrosis. In severe COVID-19, inflammatory monocyte/macrophage populations in the airways and lung are linked to NLRP3-centered inflammasome activation, IL-1β production, and pyroptotic inflammatory programs, with both infected and bystander myeloid cells contributing to pathology [9–11]. This approach addresses a key knowledge gap in translating systemic inflammasome therapies to targeted lung delivery, ensuring therapeutic benefit while minimizing off-target effects—critical for clinical development of inflammasome-based treatments for lung-specific conditions. By integrating *ex vivo* human lung perfusion and ventilation with bronchoscopic microdosing and distal airway sampling, we establish a translational platform for direct assessment of pulmonary drug delivery, cellular uptake, and pharmacology in humans. Using ADS032, a dual NLRP1/NLRP3 inflammasome inhibitor[8], as a test compound, we demonstrate the feasibility of localized intrapulmonary microdosing, quantitative spatial pharmacokinetic analysis, and cell-level target engagement in the human lung prior to Phase 1 evaluation.

## Methods

### Formulation and International Council of Harmonization (ICH) stability studies of ADS032 drug product

Formulation development and ICH stability studies for the ADS032 drug product were performed by the Healthcare Technology Accelerator Facility (HTAF), University of Edinburgh, UK. A range of excipients and vehicles were screened to develop a liquid formulation at a target concentration of 66.7 µg/mL (157 µM). A formulation containing 0.01% Tris base in water for injection (WFI) demonstrated stability for 14 days at room temperature, as assessed using a validated high-performance liquid chromatography (HPLC) method.

An ICH-compliant stability study was subsequently conducted in accordance with ICH Q1A(R2) guidelines. Stability was evaluated under multiple storage conditions (40°C, 25°C, and 2–8°C) and container orientations using internally sterile, nitrogen-flushed 10 mL Type I clear glass vials sealed with grey bromobutyl rubber stoppers. Vials were sampled at predefined intervals, and purity, concentration, and pH were measured. No significant degradation was observed over 28 months when stored upright at 25°C or 2–8°C. Evaluation of the stability data in accordance with ICH Q1E supported assignment of a 40-month shelf life from the date of manufacture.

### Manufacture of GMP-grade ADS032 sterile drug product

The sterile ADS032 drug product was manufactured to Good Manufacturing Practice (GMP) standards by the Healthcare Technology Accelerator Facility (HTAF), University of Edinburgh. Sterilisation was achieved by membrane filtration, followed by aseptic filling into sterile, closed vials within a Grade A isolator. Product quality was assured through compliance with defined and validated manufacturing procedures and testing against internal specifications, in accordance with European GMP guidance for sterile investigational medicinal products.

### Synthesis and characterisation of fluorescent ADS032–NBD

ADS032–NBD was synthesised by conjugation of ADS032 free acid to a nitrobenzoxadiazole (NBD) fluorophore via a polyethylene glycol (PEG) linker. ADS032 free acid (100 mg, 0.25 mmol, 1 eq), N-(tert-butoxycarbonyl)-2,2′-(ethylenedioxy)diethylamine (68 mg, 0.27 mmol, 1.1 eq), and 4-dimethylaminopyridine (0.3 mg, 0.03 mmol, 0.1 eq) were dissolved in anhydrous dichloromethane (5 mL). N,N′-dicyclohexylcarbodiimide (56 mg, 0.27 mmol, 1.1 eq) was added, and the reaction mixture was stirred at room temperature for 16 h. The mixture was filtered, washed with dichloromethane, and concentrated under reduced pressure.

The resulting ADS032–PEG–Boc intermediate was purified by flash chromatography (cyclohexane/ethyl acetate gradient from 50:50 to 100% ethyl acetate) to yield 80 mg of a colourless oil. Boc deprotection was performed by stirring the intermediate in a 1:1 (v/v) dichloromethane/trifluoroacetic acid mixture for 1 h at room temperature, with reaction completion monitored by thin-layer chromatography. Solvent removal under reduced pressure yielded ADS032–PEG–NH₃⁺·TFA⁻ as a white solid.

The deprotected intermediate (80 mg, 0.12 mmol, 1 eq) was dissolved in ethanol/water (3:1, v/v), and potassium carbonate (25 mg, 0.18 mmol, 1.5 eq) and 4-chloro-7-nitrobenzofurazan (NBD–Cl; 36 mg, 0.18 mmol, 1.5 eq) were added. The reaction mixture was sealed in a microwave tube and heated to 75°C for 15 minutes. Following cooling, the mixture was diluted with water (∼10 mL) and extracted three times with ethyl acetate (5 mL). The combined organic layers were dried over sodium sulfpate and concentrated under reduced pressure.

The crude product was purified by preparative HPLC and lyophilised to yield ADS032–NBD as a brown powder (44 mg, 25% overall yield over three steps).¹H NMR (500 MHz, DMSO-d₆, δ ppm): 9.44 (br s, 1H), 8.50 (d, J = 8.7 Hz, 1H), 7.82 (m, 4H), 7.64 (d, J = 8.3 Hz, 2H), 7.31 (d, J = 8.3 Hz, 2H), 6.46 (d, J = 8.7 Hz, 1H), 3.71 (t, J = 5.7 Hz, 2H), 3.64 (br s, 1H), 3.57–3.52 (m, 4H), 3.39 (t, J = 5.7 Hz, 2H), 3.17 (m, 4H), 3.06 (dd, J = 7.5 Hz, 4H), 2.60 (t, J = 7.5 Hz, 2H), 2.09 (t, J = 7.5 Hz, 2H), 1.81 (dt, J = 15.0, 7.5 Hz, 2H), 1.50 (m, 4H), 0.83 (t, J = 7.5 Hz, 2H).¹³C NMR (126 MHz, DMSO-d₆, δ ppm):171.82, 143.88, 142.37, 137.88, 135.84, 129.11, 127.41, 127.13, 126.90, 69.77, 69.49, 69.12, 49.77, 34.68, 34.18, 26.83, 21.73, 10.99. Full synthetic procedures and analytical characterisation are provided in the Supplementary Methods.

### Human alveolar macrophage isolation from *ex vivo* human lungs

The *ex vivo* human lung ventilation protocol has been described previously [17]. Briefly, human lungs were ventilated with a tidal volume of 7 mL/kg of ideal body weight at a respiratory rate of 12 breaths per minute[12]. A bronchoscope (Ambu® aScope™ 4 Broncho (Regular 5.0/2.2) was wedged into a lobar subdivision, and bronchoalveolar lavage (BAL) was performed by instilling 200 mL of 0.9% sodium chloride, which was then aspirated.

BAL fluid was filtered through a cell strainer and centrifuged at 300 × g for 5 minutes at 4°C. The resulting cell pellet was resuspended in a total volume of 10 mL phosphate-buffered saline (PBS). The filtrate was centrifuged and washed up to five times using the same volume of PBS supplemented with antibiotics (50 IU/mL penicillin and 100 μg/mL streptomycin; prepared as 5 mL penicillin/streptomycin in 500 mL PBS).

Cells were counted and adjusted to a concentration of 2 × 10⁶ cells/mL in alveolar macrophage (AM) culture medium (Iscove’s Modified Dulbecco’s Medium supplemented with 1% penicillin/streptomycin, 1% L-glutamine and 10% fetal bovine serum. Aliquots of 0.5 mL cell suspension were seeded into 24-well plates and incubated for 1 hour at 37°C to allow macrophage adherence. Non-adherent cells were removed by washing with warm PBS, and fresh culture medium was added.

### Inflammasome activation and inhibition

Experiments were performed on cultured primary human alveolar macrophages (AMs) plated at a density of 0.5–2 × 10⁶ cells per well. Cells were primed with *Escherichia coli* lipopolysaccharide B5 (LPS; final concentration 100 ng/mL) diluted in culture medium for 2 hours. Following LPS priming, media were aspirated and replaced with serum- and BSA-free medium containing inflammasome inhibitors (MCC950 at 1μM or ADS032 at concentrations of 1, 2, 50, 100 and 350μM), which were applied for 1 hour. Nigericin was then added to each well (final concentration 3.35 μM) to induce inflammasome activation. After 2 hours, supernatants were collected and cells were snap-frozen in the culture wells for downstream analysis. This protocol was adapted from previously described methods [18]. MCC950 was used as a commercially available inhibitor of the inflammasome pathway[13].

### Cell imaging

BAL cells were imaged using an EVOS M5000 imaging system microscope.

### Cytokine analysis

Cell culture supernatants were snap-frozen on dry ice immediately after collection and stored at −80°C until analysis. Cytokine concentrations were quantified using a four-plex immunoassay (IL-1β, TNF, IL-6, and IL-8) on the ELLA platform (ProteinSimple, Bio-Techne, USA), according to the manufacturer’s instructions.

### Flow cytometry

Antibody staining cocktails were prepared in fluorescence-activated cell sorting (FACS) staining buffer consisting of phosphate-buffered saline (PBS) supplemented with 2% flow cytometry staining buffer (Gibco) and Brilliant Violet Plus buffer (BD Biosciences, 566385). Samples were incubated with 50μL of antibody cocktail for 20 minutes at room temperature in the dark. Following staining, samples were washed and fixed before analysed on a Cytek Aurora spectral flow cytometer.

### Lung tissue digestion from *ex vivo* human lungs

Lung tissue was transferred to a Petri dish or bijoux containing 1 mL Dulbecco’s Modified Eagle Medium (DMEM) and mechanically minced using sterile scissors until finely dissociated. The tissue was then transferred to a 50 mL conical tube and topped up to a final volume of 20 mL with DMEM. Samples were centrifuged at 350 × g for 5 minutes at room temperature, and the supernatant was discarded.

The resulting pellet was resuspended in 5 mL pre-warmed DMEM, followed by the addition of 5 mL pre-warmed collagenase (Merck)/DNase (Merck) digestion mix to achieve final concentrations of 1 mg/mL collagenase I, 1 mg/mL collagenase IV, and 0.1 mg/mL DNase. Samples were incubated for 1 hour at 37°C with agitation.

Following digestion, the cell suspension was filtered through a 70 μm cell strainer into a 50 mL conical tube, with residual tissue washed through the strainer using DMEM. The filtrate was centrifuged at 350 × g for 5 minutes at room temperature, and the supernatant was removed. Red blood cells were lysed by adding an appropriate volume (1–10 mL) of red blood cell lysis buffer and incubating for 10 minutes at room temperature. Cells were then centrifuged at 350 × g for 5 minutes at room temperature, the supernatant discarded, and the final cell pellet resuspended for cell counting and downstream applications.

### *Ex vivo* human lung microdose experiment

Human lungs were separated into right and left lobes. A perfusate solution was prepared consisting of 1 L high-glucose Dulbecco’s Modified Eagle Medium (DMEM), 70 g bovine serum albumin (BSA), 5 g Dextran-40, and 5,000 international units of heparin. An endotracheal tube was inserted into the trachea or residual main bronchus, and a pulmonary artery catheter was sutured into the remaining pulmonary arteries. The right lung was perfused and ventilated according to an established *ex vivo* lung perfusion (EVLP) protocol[14] using a Dräger Savina 300 ventilator. The remaining lung was reserved for follow-up experiments.

A baseline 2 mL perfusate sample was collected from the circuit and immediately snap-frozen on dry ice and stored at −80°C. The distal alveolar lung space was visualised using a fibre-based confocal endomicroscopy system as previously described. ADS032 was administered intra-alveolarly (1.5 mL; 100 μg) via the delivery fibre (Panoptes) . To label the area for further evaluation, up to 1ml of 2 mM methylene blue was administered to demarcate the region.

Perfusate samples (2 mL) were collected from the circuit at predefined time points and immediately snap-frozen on dry ice and stored at −80°C. Microlavage samples from the distal lung were collected at corresponding time points using an Erbe catheter after 1mL of saline delivery.

### Liquid chromatography–mass spectrometry

For full Liquid Chromatography Mass spectrometry Materials and Methods see supplemental methods. Briefly, quantitative analysis was performed using a Sciex QTRAP 6500+ tandem quadrupole mass spectrometer operated in positive multiple reaction monitoring (MRM) mode, coupled to a Waters Acquity I-Class UPLC system. Chromatographic separation was achieved using a Waters BEH C18 column (50 × 2.1 mm, 1.7 μm particle size) at a flow rate of 0.4 mL/minutes. The mobile phases consisted of water with 0.1% formic acid (A) and acetonitrile with 0.1% formic acid (B), with a total gradient run time of 6 minutes.

### Aspirate samples from experiment with *ex vivo* human lungs

ADS032 and the internal standard, d9-progesterone, were extracted from aspirate samples alongside a calibration curve using a phospholipid depletion plate (Biotage PLD+) to remove phospholipids and proteins. Sample preparation was performed using a Biotage Extrahera automated pipetting and extraction system operating under positive pressure. Extracts were dried using a Biotage SPE Dry 96 system and subsequently resuspended in the appropriate solvent prior to LC–MS/MS analysis.

### Lung tissue samples from *ex vivo* human lungs

Approximately 50 mg of lung tissue was weighed and transferred to screw-cap tubes containing 2.8 mm ceramic beads. Acetonitrile containing 0.1% formic acid (0.5 mL) and the internal standard were added, and samples were homogenised using an Omni Bead Ruptor system. Homogenates were centrifuged, and the resulting supernatants were passed through a Biotage Filter+ (0.2 μm) filter plate under positive pressure using a Waters Pressure+ 96 unit. The filtered samples were then processed through a Biotage PLD+ plate under positive pressure to remove residual phospholipids and proteins. Eluates were collected, dried using the Biotage SPE Dry 96 system, and resuspended for LC–MS/MS analysis.

### Intratarget Microdosing Trial (Micro) trial design and participants

Micro was a single-centre, open-label, Phase 0 intratarget microdosing study. Participants were recruited between November 2023 and December 2024. Eligible participants were adults (≥18 years) with suspected or confirmed interstitial lung disease (ILD) or bronchiectasis who were scheduled to undergo bronchoscopy as part of standard clinical care. All participants provided written informed consent.

Exclusion criteria included pregnancy or breastfeeding, known hypersensitivity to ADS032, enrolment in a concurrent clinical trial of an investigational medicinal product, receipt of systemic corticosteroids or other immunomodulatory therapies (including azathioprine, mycophenolate, ongoing chemotherapy, or macrolide antibiotics), and any condition that, in the investigator’s judgement, would preclude safe completion of bronchoscopy or study procedures.

Given the exploratory nature of this Phase 0 study, no formal sample size calculation was performed, and participants were not randomised.

### Interventions

Participants received a single intrapulmonary microdose of ADS032 (93ug/ 1.5mL) administered bronchoscopically to a distal lung segment using a medically approved catheter. An equivalent volume of 0.9% sodium chloride was administered to the contralateral lung segment, which served as an internal control. Bronchoscopic procedures were conducted according to standard local practice and British Thoracic Society guidelines.

Topical local anaesthesia was applied to the oropharynx, with conscious sedation achieved using intravenous midazolam and/or a short-acting opioid. Bronchoscopic sampling was initiated 10 minutes following ADS032 administration. Sampling modalities included bronchoalveolar lavage, distal airway microlavage via flexible catheter, bronchoabsorption, and bronchial brushings, performed in both treated and control lung segments.

Peripheral blood samples were collected immediately prior to bronchoscopy and at 30 minutes, 1 hour, and 4 hours post-procedure. Blood tubes were centrifuged at 1400g for 10 minutes at 4℃ and supernatant was collected as plasma or serum which was stored in 1mL tubes at -80℃ before analysis.

Protocol refinements were introduced after participant 4 following interim analysis of the initial 4 patients to minimise the risk of cross-contamination between treated and control lung segments. The order of administration of ADS032 and control saline was altered with the scope removed from the participant’s airway completely with an intra procedural flush. The full trial protocol is provided in the Supplementary Materials.

### Outcomes

The primary outcome was early mechanistic evaluation of ADS032 through quantification of inflammasome-associated biomarkers (IL-1β, TNF, IL-6, IL-18, and ATP) in bronchoalveolar lavage fluid.

Secondary outcomes included assessment of ADS032 pharmacokinetics in plasma samples collected up to 4 hours post-administration, quantified using liquid chromatography–mass spectrometry.

Exploratory outcomes included analysis of cytokines, chemokines, and cell surface markers of target engagement in respiratory samples, as well as quantification of ADS032 concentrations in bronchoalveolar lavage fluid, distal airway microlavage, recovered airway cells.

### Trial registration

The study was registered with the ISRCTN clinical trial registry (ISRCTN35867933).

#### Micro trial sample processing

BAL sample were passed through a 40um cell strainer to remove debris before centrifuge for 10 minutes at 300g in a pre-chilled (4℃) centrifuge. BAL supernatant was removed, aliquoted and snap-freeze on dry ice. BAL cells remaining re-suspended in 10mL cold PBS, transferred into micro tubes then pelleted again and snap-freeze as cell pellets. For distal airway microlavage samples, the samples were pooled first, then evenly aliquoted into 3 vials before snap-freeze on dry ice. Bronchial brushings were kept in cold PBS, cells were removed from brushes by vortexing for 1 minute and then rinsed with cold PBS. Brush cells were then pelleted by centrifugation in a pre-chilled centrifuge at 300g for 10 minutes. For ADS032 extraction from BAL and brush cells, a cell pellet was retrieved from biobank, defrosted, then lysed with ultra-pure water. After 10 minutes incubation cell debris was removed by centrifugation at 1400g at 4℃ for 10 minutes, then supernatant was collected for mass-spec analysis. All samples were transferred and stored in -80℃ before analysed by per protocol.

### ATP/AMP measurement

Samples were filtered using 0.2um PTFE Captiva syringe filters (Agilent) and stored at -20C. Sample preparation to be described here.

Chromatographic analyses were performed using an Agilent High-Pressure Liquid Chromatography system 1200 series equipped with a quaternary pump, an autosampler, a column oven and a variable wavelength detector (Agilent Technologies, Santa Clara, CA, USA). The system control and data processing were performed using an OpenLab CDS LC ChemStation from Agilent. Separation was carried out on a reverse-phase Phenomenex XB-C18 column (2.1 × 150 mm, 2.6 µm) maintained at 25 °C. The injection volume was 5 µL. The compounds in standard and sample solutions were separated using a gradient (see table below) between the mobile phases of 50 mM Potassium DiHydrogen Phosphate in Water (mobile phase C), 0.1% Formic Acid in Water (mobile phase A) and 0.1% Formic Acid in Acetonitrile (mobile phase B). The absorbance was monitored at 254 nm.

## Results

### ADS032 inhibits inflammasome activation in primary human alveolar macrophages

Figure 1A shows the chemical structure of ADS032 and the addition of a nitrobenzoxadiazole (NBD) moiety to generate a fluorescently labelled compound. To confirm inflammasome inhibition in primary human cells, alveolar macrophages (AMs) were isolated from bronchoalveolar lavage fluid obtained from *ex vivo* human lungs. Inflammasome activation with lipopolysaccharide (LPS) and nigericin resulted in robust IL-1β release, which was completely inhibited by the prototypic NLRP3 inhibitor MCC950 at concentrations of 1 μM and 100 nM (Fig. 1B), consistent with its use as a positive control for inflammasome inhibition [18].

**Figure 1:**
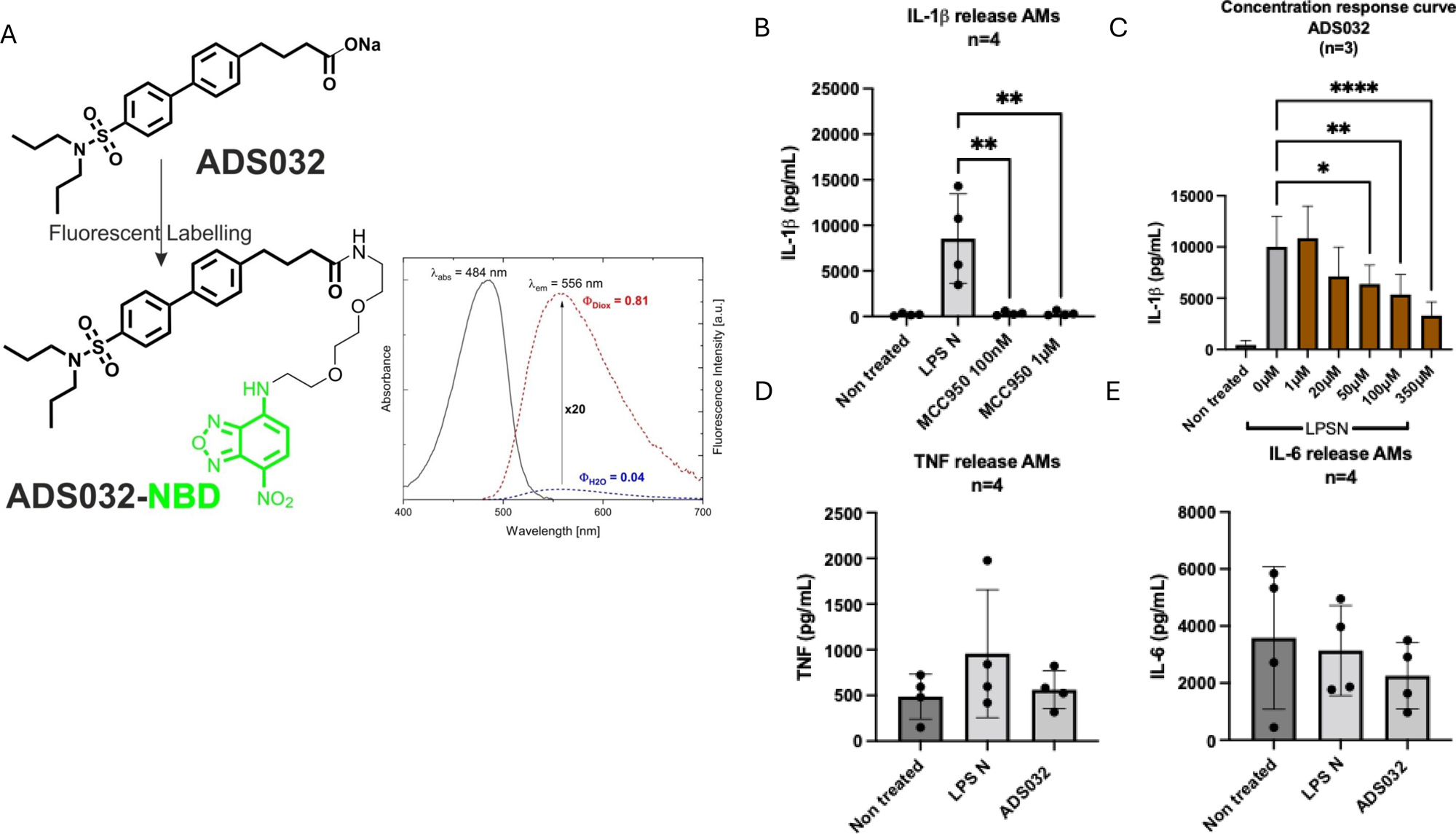
ADS032 structure and inflammasome inhibition. Chemical structure and effect of inflammasome activation/ inhibition on human alveolar macrophages (AM) which have been retrieved from ex vivo human lungs. Inflammasome activation via 100ng/mL LPS + nigericin 3.35μM (LPS N). Inhibition by MCC950 and ADS032. Analysis by 1 way ANOVA. Mean and standard deviation plotted and p values plotted where <0.005. A- Chemical structure of ADS032 + fluorescence NBD probe. B - IL-1β release from AMs is inhibited by the addition of MCC950 (commercially available inflammasome inhibitor). C-concentration response curve of effect of ADS032 on IL-1β release from AMs. D & E- TNF and IL-6 release from AMs is not affected by activation via LPS+ nigericin or inhibition with ADS032 at a concentration of 100µM. N=4 with 2 technical repeats for each experiment. Fig 1C N= 3 with 2 technical repeats for each experiment.

Importantly ADS032 significantly inhibited IL-1β release from human AMs following NLPR3 inflammasome stimulation, with significant inhibition observed at concentrations ≥50 μM (Fig. 1C), demonstrating a concentration-dependent effect consistent with previous reports[8]. In contrast, ADS032 did not significantly inhibit TNF or IL-6 release under the same conditions (Fig. 1D, E), supporting selective modulation of the inflammasome pathway rather than broader suppression of inflammatory signalling.

### Rapid cellular uptake of ADS032 visualised in *ex vivo* human lungs

ADS032-NBD labelled AMs as seen in Fig 2A. During *ex vivo* human lung perfusion and ventilation experiments, ADS032–NBD was administered directly to the distal alveolar space under real-time visualization using the alveolar endomicroscopy platform. Cellular uptake of ADS032–NBD was observed within minutes of exposure. *In vitro* treatment of isolated human AMs with ADS032–NBD resulted in substantial inhibition of IL-1β release (Supp Fig. 4), although inhibition was reduced compared with unlabelled ADS032, consistent with partial attenuation of potency following fluorophore conjugation.

**Figure 2:**
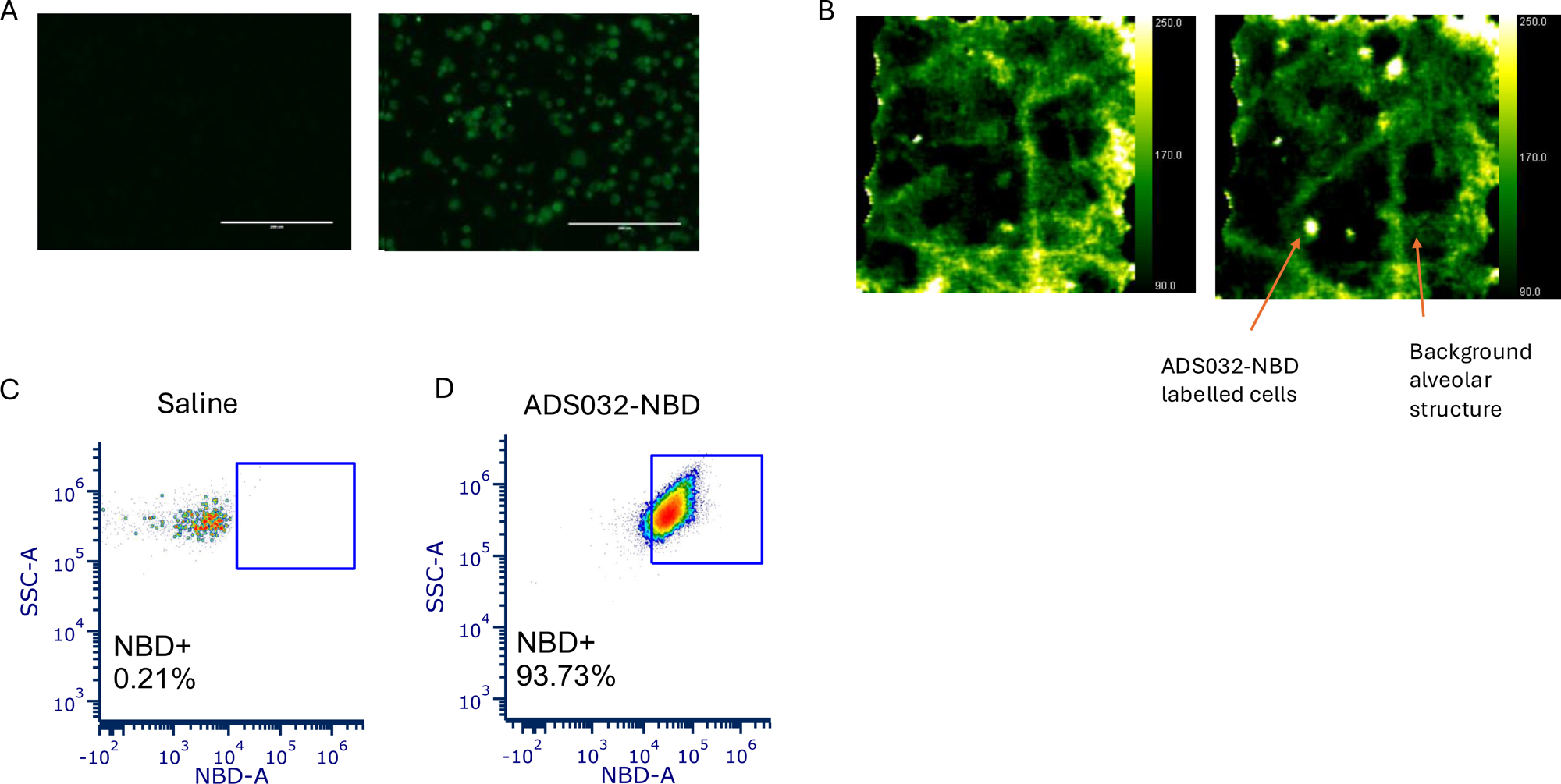
Ex vivo uptake and alveolar endomicroscopy. Fluorescently labelled ADS032 allows for visualisation and confirmation of cellular drug uptake. A- EVOS microscope images of AMs – untreated (left image) and treated with ADS032-NBD (right image). Scale bar 400µm. Cells exposed to ADS032-NBD have increased green fluorescence. B-ADS032-NBD retains a degree of inhibitory ability. IL-1β release from AMs. C- Representative image of alveolar endomicroscopy following delivery of microdose (0.5-1.5mLs) ADS032-NBD into the alveolar space of ventilated ex vivo lungs. Bright spots with high green intensity are presumably labelled alveolar inflammatory cells. Images taken with Kronoscan imaging system using Panoptes imaging/ delivery fibre. D & E- Flow cytometric analysis of digested lung tissue following administration of microdose (1.5mLs) of ADS032-NBD to ventilated alveolar space demonstrating NBD shift. Plots display cells with macrophage phenotype (CD45/ Lin-/ CD3-/ CD11b+, HLADR+). E- area of digested lung + normal saline control.

Alveolar endomicroscopy enabled direct visualisation of NBD-positive inflammatory cells within ventilated alveoli (Fig. 2B), with preserved alveolar architecture visible in the background. Cellular uptake was further confirmed by flow cytometric analysis following digestion of lung tissue exposed to ADS032–NBD. Lung regions receiving ADS032–NBD demonstrated a clear fluorescence shift compared with saline-treated control regions (Fig. 2C, D). Macrophage populations were identified as CD45⁺Lin⁻CD3-CD11b+HLA-DR⁺ cells, and a distinct NBD shift was observed only in drug-exposed tissue, confirming spatially restricted cellular uptake.

### Cell-type–specific uptake of ADS032 in the *ex vivo* lung

To enable more detailed phenotyping of drug uptake, a higher-volume intrapulmonary delivery experiment was performed in ventilated *ex vivo* lungs. A total of 100 mL solution containing ADS032–NBD (90 mL saline plus 10 mL of 100 μM ADS032–NBD) was administered endobronchially, followed by bronchoalveolar lavage and lung tissue digestion after 10 minutes. A contralateral control lobe received saline alone.

Flow cytometric analysis demonstrated uptake of ADS032–NBD across multiple cell populations (Fig. 3). Strong fluorescence shifts were observed in monocyte and macrophage populations, particularly alveolar macrophages expressing CD206 and CD163 (Fig. 3A–C). Uptake was also detected in neutrophils (live CD45⁺Lin⁺CD11b⁺ cells; Fig. 3F) as well as lymphocyte populations, including CD3⁺ T cells (Fig. 3D). NBD shift was observed in epithelial cells following lung tissue digestion and cells recovered by lavage (Fig. 3E). The NBD shift seen in all cell populations is suggestive that ADS032 is not selective in its uptake to any particular cell type but the results overall confirm the ability of NBD conjugation to confirm compound uptake following bronchoscopic administration.

**Figure 3:**
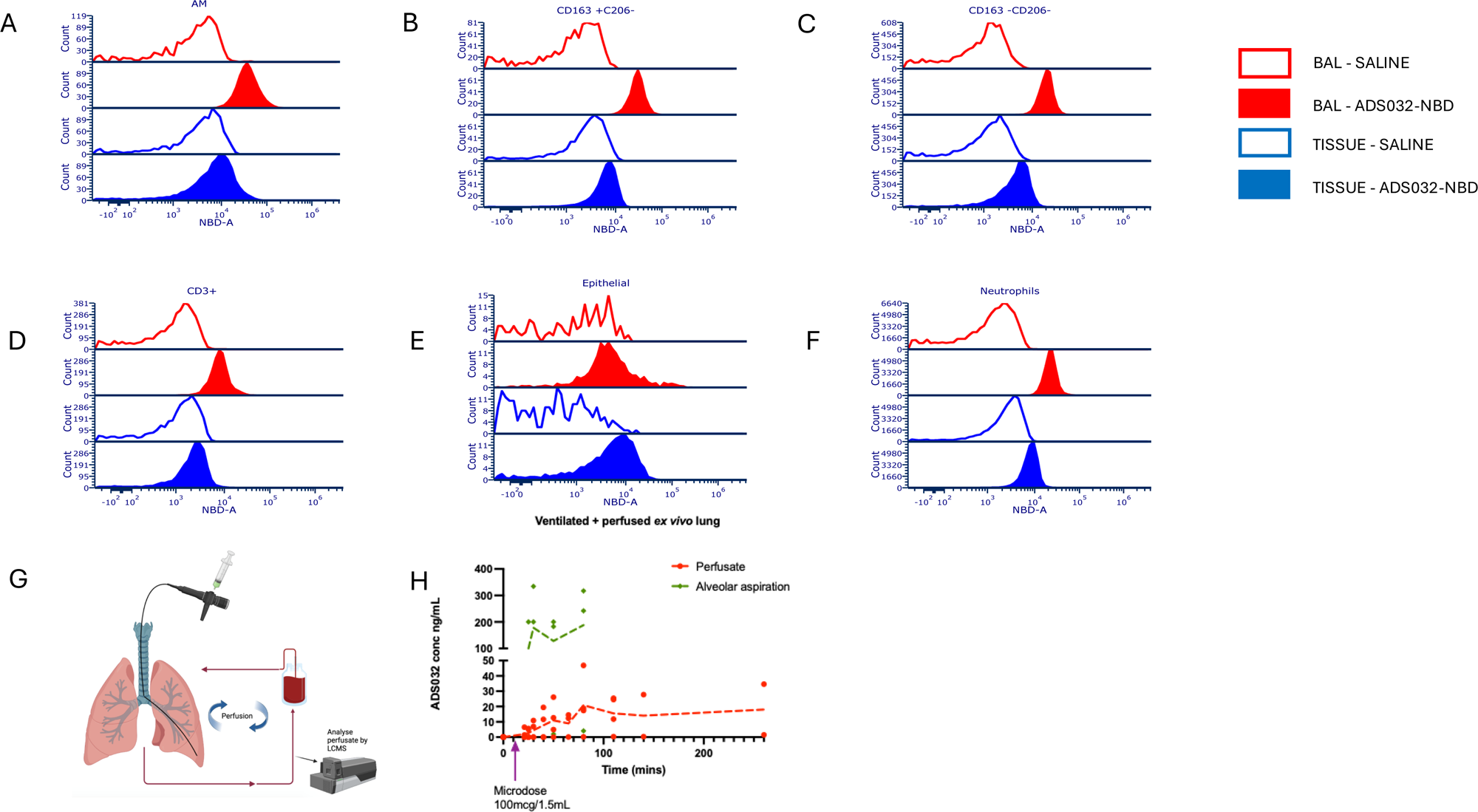
Ex vivo uptake and alveolar endomicroscopy. Fluorescently labelled ADS032 allows for visualisation and confirmation of cellular drug uptake. A- EVOS microscope images of AMs – untreated (left image) and treated with ADS032-NBD (right image). Scale bar 400µm. Cells exposed to ADS032-NBD have increased green fluorescence. B-ADS032-NBD retains a degree of inhibitory ability. IL-1β release from AMs. C- Representative image of alveolar endomicroscopy following delivery of microdose (0.5-1.5mLs) ADS032-NBD into the alveolar space of ventilated ex vivo lungs. Bright spots with high green intensity are presumably labelled alveolar inflammatory cells. Images taken with Kronoscan imaging system using Panoptes imaging/ delivery fibre. D & E- Flow cytometric analysis of digested lung tissue following administration of microdose (1.5mLs) of ADS032-NBD to ventilated alveolar space demonstrating NBD shift. Plots display cells with macrophage phenotype (CD45/ Lin-/ CD3-/ CD11b+, HLADR+). E- area of digested lung + normal saline control.

**Figure 4:**
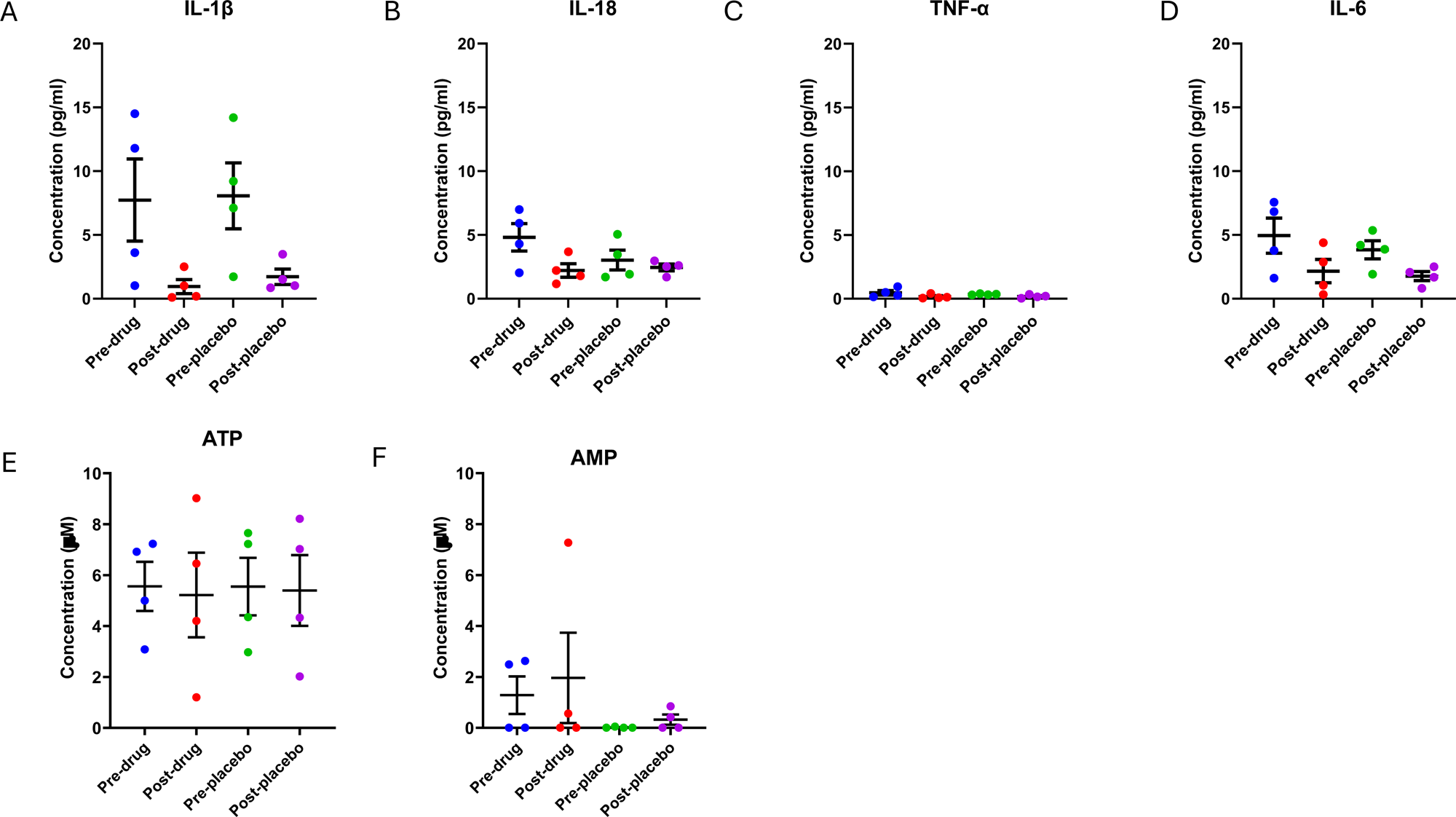
Clinical cytokine readouts. Cytokine levels in bronchoalveolar lavage fluid, analysed by ELLA system. Results analysed by 2 way ANOVA. For each patient 4 BALs collected during the bronchoscopic procedure; pre- drug and pre- placebo (both baselines) and post 0.9% sodium chloride placebo and post ADS032. The post BALs were collected 10 mins after placebo/ ADS032 installation. For all results only participants 5 to 8 results analysed. A Il-1β, B IL-18, C TNF-⍺, D IL-6, E ATP (adenosine triphosphate), F AMP (adenosine monophosphate).

Taken together, these results clearly demonstrate that alveolar microdosing with ADS032 induces rapid uptake into the pulmonary microenvironment allowing effective targeting of the NLRP3 inflammasome *in situ*.

### *Ex vivo* lung perfusion enables spatial pharmacokinetic analysis

The *ex vivo* lung perfusion and ventilation system provides an excellent to model intrapulmonary pharmacokinetics under physiological conditions. A microdose of ADS032 (100 μg in 1.5 mL saline), matching the planned clinical dose, was administered endobronchially to the right middle lobe (experimental setup shown in Fig. 3G). Sequential samples of lung perfusate and distal airway microlavage were collected over time and analysed by LC–MS.

ADS032 was detectable in both alveolar aspirates and lung perfusate, enabling simultaneous assessment of local pulmonary and systemic drug exposure (Fig. 3H). Perfusate concentrations remained relatively stable for up to 4 hours following administration (concentration range 0.0-46.9 ng/mL), although this likely overestimates systemic exposure compared with the *in vivo* setting, where renal and hepatic clearance would be expected. Detection of ADS032 in low-volume alveolar microlavages (typically ∼100 μL returned following instillation of 0.5–1.0 mL saline) demonstrates the sensitivity of the analytical approach and feasibility of distal airway pharmacokinetic sampling. These findings informed and were subsequently replicated in the clinical microdosing study.

### Clinical feasibility and patient characteristics

Twelve patients were enrolled in the Micro trial, all of whom had suspected or confirmed interstitial lung disease. Patient demographics are summarised in Table 1. The mean age was 68.1 years, with 10 male and 2 female participants. Supplementary Table 1 summarises bronchoscopic sampling metrics. The mean cell yield from bronchoalveolar lavage was 1.82 million on average with all BALs (n=12) from all 4 groups with median recovery of 1.2 million, minimum 0.1 million and maximum 6.5 million. The average volume returned from distal airway microaspiration was1.4 mL on average with all microlavages (n=7, subs 5-12) from all 4 groups with median recovery of 1.485 mL, minimum 90uL and maximum 5.0 mL with all 3 aliquots been taken for each procedure.

**Table 1.** Patient demographics for patients recruited to Micro study.

| Subject number | Sex | Age Range | Diagnosis | Respiratory diagnosis subtype | Smoking status | Co-morbidities |
| --- | --- | --- | --- | --- | --- | --- |
| 1 | Female | 56-60 | ILD | Systemic sclerosis associated ILD | Non-smoker | Systemic sclerosis |
| 2 | Male | 76-80 | ILD | IPF | Ex-smoker | IHD, heart failure, pacemaker for heart block, hypertension, Raynaud's syndrome, renal stones |
| 3 | Male | 56-60 | ILD | IIP with fibrosis | Ex-smoker | Essential tremor |
| 4 | Male | 66-70 | ILD | IPF | Ex-smoker | Aortic stenosis, lung nodules, NAFLD, Raynaud's syndrome, gout, hyperlipidaemia |
| 5 | Male | 71-75 | ILD | IIP with fibrosis | Non-smoker | Atrial fibrillation, previous stroke |
| 6 | Male | 46-50 | ILD | IIP | Non-smoker | Femoral vein thrombosis, lung nodules |
| 7 | Male | 66-70 | ILD | IIP with fibrosis | Ex-smoker | COPD, hypertension, Hodgkin's lymphoma, Factor V Leiden |
| 8 | Male | 61-65 | ILD | IIP with fibrosis | Non-smoker | Nil |
| 9 | Male | 76-80 | ILD | IIP with fibrosis | Non-smoker | Hypertension, Probable IPF, Prostate Cancer 2013 - Prostatectomy |
| 10 | Female | 81-85 | ILD | IIP with fibrosis | Ex-Smoker | Hypertension, Stage 3 Chronic Kidney Disease |
| 11 | Male | 66-70 | ILD | IIP with fibrosis | Ex-Smoker | Hypertension, Diabetes, Bladder Cancer, AF, Hypercholesterolaemia, COPD, Erythrocytosis, Moderate coronary artery calcification |
| 12 | Male | 76-80 | ILD | IIP with fibrosis | Ex-Smoker | Emphysema, skin squamous cell cancer 2023, Vertigo |

Intrapulmonary microdosing is safe and permits spatially resolved pharmacokinetics in humans. There were no recorded adverse events in the Micro trial.

Key inflammasome-associated cytokines were measured in bronchoalveolar lavage fluid collected before and after ADS032 or saline instillation (Fig. 4A–F). Due to identified cross contamination in participants 1-4, only participants 5–12 are shown in the main analysis. Critically no significant differences were observed in IL-1β, TNF, IL-6, IL-18 or ATP levels when comparing pre- versus post-instillation samples or treated versus control lung segments, highlighting that ADS032 exposure does not induce an acute inflammatory response following administration ADS032 pharmacokinetics were further quantified using LC–MS in plasma and respiratory samples. Plasma ADS032 concentrations peaked at 30 minutes post-administration and remained detectable at 1 hour in most participants (Fig. 5A). By 4 hours, plasma concentrations were below the limit of detection in all but two participants. ADS032 was detectable in post-administration BAL fluid (Supplementary Fig. 1A); however, similar concentrations were observed in control lobe BAL, consistent with equipment-related cross-contamination. Accordingly, intracellular ADS032 concentrations in BAL-derived cells were comparable between treated and control lobes (Supplementary Fig. 1B).

**Figure 5:**
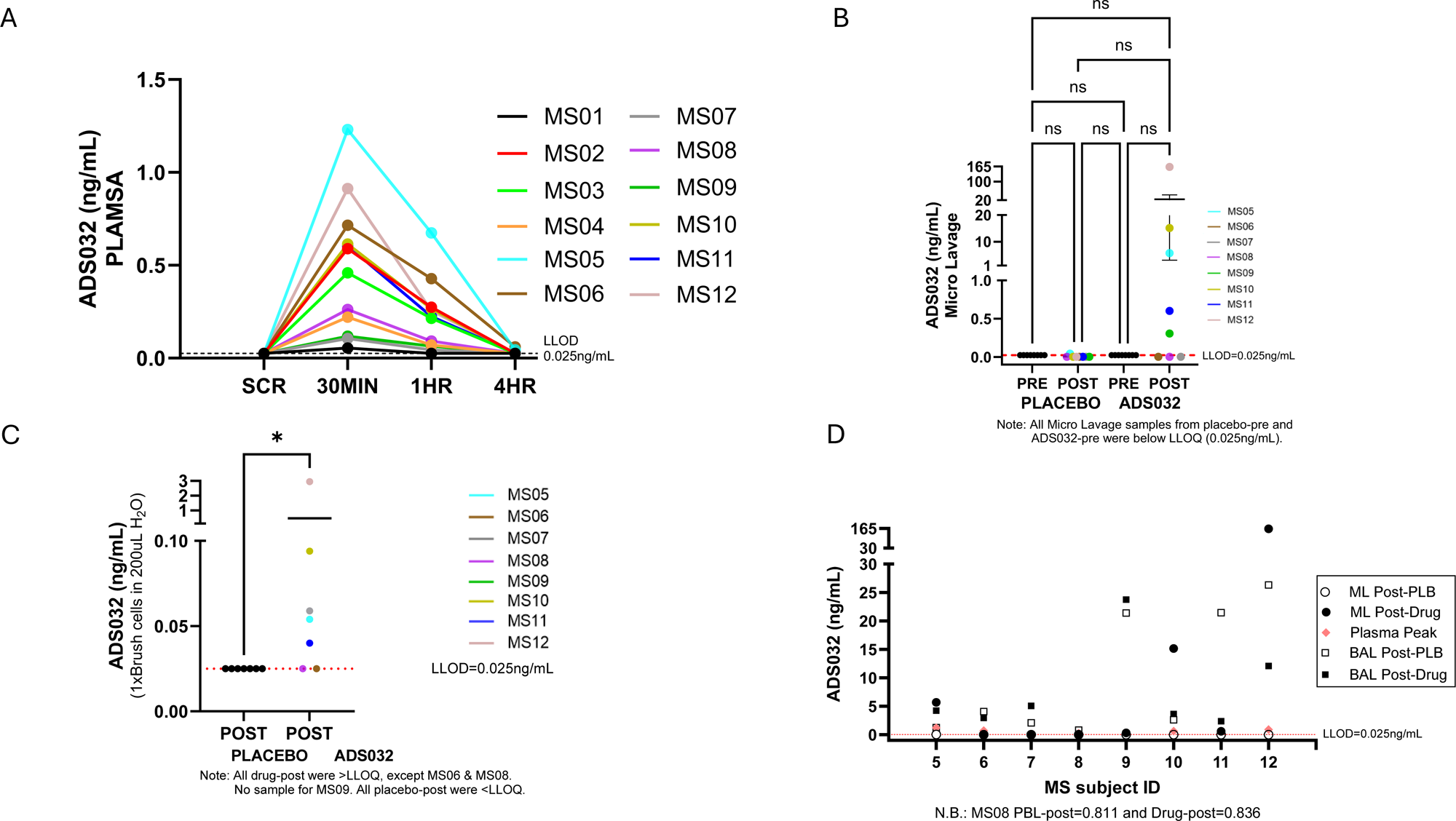
Human pharmacokinetics and spatial sampling. ADS032 concentration as measured by LC-MS. A: ADS032 concentration in blood plasma. Plasma levels checked at screening and 30 mins, 1 hour and 4 hours post administration. At screening (pre procedure) blood sampling all subjects were below the below limit of quantification (LLOQ) 0.025ng/mL. Peak concentration at 30 minutes, with drop at 1 hour. At 4 hours post administration all subjects were below LLOQ, except MS05 and MS06. B: ADS032 concentration in microlavage samples, taken with use of flexible catheter passed through working channel of bronchoscope. All baseline pre placebo and pre ADS032 samples were below the LLOQ. Statistically significant rise in concentration of ADS032 in microlavage sample from post ADS032 lobe. C: ADS032 concentration in cells obtained through distal brushings, taken with use of cytology brush passed through working channel of bronchoscope. Only samples from post placebo and post ADS032 lobes were obtained. All placebo lobe samples were below LLOQ. ADS032 was detectable in all but 2 samples from the post ADS032 lobe (MS06 and MS08). No sample obtained for MS09 due to issues during bronchoscopic procedure.

In contrast, no contamination was evident in distal airway microlavage samples. ADS032 was detected in five treated-lobe microlavages at a mean concentration of 36.923 ng/mL (min 0.306ng/mL, max 162.874 ng/mL and median 5.69 ng/mL. ADS032 was only detected in one control microlavage (Participant 5: 0.0430 ng/mL) (Fig. 5B). Similarly, distal bronchial brushings revealed detectable ADS032 in five treated lobes, with no detectable drug in control lobes (Fig. 5C). These findings indicate that distal airway microaspiration and brushing provide superior spatial resolution for intrapulmonary pharmacokinetic assessment compared with bronchoalveolar lavage.

An overview of ADS032 concentrations measured across plasma, BAL, and microlavage samples for participants 5–12 is shown in Fig. 5D

## Discussion

High attrition rates in drug development remain a major challenge for global public health and healthcare sustainability. Despite sustained efforts to improve efficiency, most compounds entering clinical development fail in Phase II or Phase III trials, predominantly because of insufficient clinical efficacy rather than unanticipated toxicity[15–17]. These late-stage failures highlight a persistent gap between preclinical models and human disease biology and underscore the need for approaches that enable earlier, human-relevant assessment of pharmacokinetics, target engagement, and mechanism of action.

Phase 0 clinical trials, particularly intratarget microdosing (ITM) strategies, offer a framework to address this gap. By delivering subtherapeutic doses directly to the target tissue, ITM enables local drug exposure, spatially resolved sampling, and mechanistic interrogation while maintaining wide systemic safety margins. Importantly, Phase 0 and Phase I trials can be conducted in parallel, allowing compounds to be prioritised or deprioritised based on early human data rather than prolonged preclinical extrapolation.

In respiratory medicine, the limitations of traditional animal models are increasingly recognised. Differences in lung size, anatomy, immune composition, and disease pathophysiology limit the predictive value of rodent studies and contribute to poor translation of apparently promising anti-inflammatory therapies. Human-relevant platforms—including *ex vivo* lung perfusion and ventilation systems and early in-human evaluation—therefore represent a necessary evolution in pulmonary drug development, enabling direct interrogation of drug behaviour in intact human lung tissue.

Using ADS032 as a tool compound, we demonstrate the feasibility of an integrated translational pipeline spanning *ex vivo* human lungs and a first-in-human intrapulmonary microdosing study. ADS032 exhibited favourable early characteristics, including rapid uptake by alveolar macrophages, selective inhibition of inflammasome-mediated IL-1β release in stimulated primary human macrophages, and no evidence of toxicity in preclinical or cellular assays[8]. In both *ex vivo* lungs and patients, ADS032 rapidly translocated from the distal airways into the perfusate or systemic circulation, providing early pharmacokinetic data to inform subsequent dose modelling and clinical development. Ventilated *ex vivo* lung experiments demonstrated excellent uptake of ADS032 into innate myeloid cells (including alveolar macrophages), which is highly favorable for targeting NLRP3-driven inflammation. Good uptake was also observed in T and B lymphocytes, while epithelial cell uptake was detectable but more modest. These preliminary findings are encouraging with respect to dual NLRP1/NLRP3 coverage and support further progression toward Phase I evaluation.

Collectively, these human-relevant findings with ADS032—a novel dual NLRP1/NLRP3 inhibitor—offer highly encouraging prospects for successful and safe advancement into Phase I clinical trials.

The demonstrated rapid cellular uptake (particularly in myeloid cells relevant to NLRP3), detectable epithelial engagement (supportive of NLRP1 targeting), favorable spatial pharmacokinetics, and absence of acute safety signals substantially de-risk progression beyond this exploratory Phase 0 study. These attributes warrant continued investment to enable full clinical evaluation in inflammatory lung diseases where inflammasome dysregulation plays a central role.

We did not observe significant differences in bronchoalveolar lavage cytokine concentrations between treated and control lung segments. This outcome is consistent with the study design and dose intent: the administered microdose was not expected to elicit robust pharmacodynamic effects detectable by bulk lavage, and BAL itself lacks the spatial resolution required for intratarget microdosing studies. Indeed, comparable ADS032 concentrations in treated and control lobe BAL fluid indicate procedural cross-contamination, likely related to bronchoscopy dynamics such as coughing or fluid redistribution. Importantly, early findings prompted protocol refinement, reducing contamination in later participants and illustrating the adaptive value of early-phase ITM studies.

In contrast, distal airway microlavage and targeted bronchial brushings provided markedly improved spatial resolution. Using these techniques, ADS032 was detectable more frequently in treated lung segments, both in fluid samples and exclusively in retrieved brushed cells, with no evidence of contamination in control regions. Notably, intracellular drug uptake was measurable despite administration of a subtherapeutic microdose, demonstrating the sensitivity and translational potential of these approaches. These findings suggest that distal sampling strategies are more appropriate than BAL for pharmacokinetic and mechanistic interrogation in intrapulmonary microdosing studies.

Several limitations warrant consideration. This was a small, heterogeneous Phase 0 study not designed to assess clinical efficacy or disease-specific pharmacodynamics. The *ex vivo* lung perfusion model, while highly translational, does not fully recapitulate systemic clearance, immune trafficking, or chronic inflammatory signalling present *in vivo*[18]. Its utility has been described in development of drugs for nebulisation [19]. Fluorescent labelling of ADS032 reduced compound potency, limiting quantitative extrapolation from uptake studies. These limitations are inherent to early-phase intratarget microdosing and reinforce the role of this approach as a platform for early human pharmacology rather than definitive efficacy assessment.

In summary, this study provides a first-in-human demonstration of intrapulmonary intratarget microdosing integrated with *ex vivo* human lung modelling. This approach enables direct evaluation of pulmonary drug delivery, cellular uptake, and spatial pharmacokinetics in humans before Phase I trials. Future studies will apply this platform to larger patient cohorts, incorporate high-resolution proteomic and transcriptomic analyses of distal airway samples, and evaluate therapeutic candidates under conditions of active pathway engagement. More broadly, intrapulmonary intratarget microdosing represents a scalable and adaptable framework for accelerating development of lung-directed therapies and reducing late-stage clinical attrition.

## Data Availability

All data produced in the present study are available upon reasonable request to the authors

## Declarations

### Ethics approval and consent to participate

The Micro study received favourable ethical approval from the South Central Oxford B Research Ethics Committee and NHS Lothian Research and Development. The Medicines and Healthcare products Regulatory Agency determined that the study was not a Clinical Trial of an Investigational Medicinal Product and did not require MHRA oversight.

### Consent for publication

Not applicable.

### Availability of data and materials

Data are available from the corresponding author upon reasonable request.

### Authors’ contributions

All authors contributed to study design, data acquisition, analysis, and manuscript preparation.

## Acknowledgements

We thank the study participants and their families for their generous donations to the advancement of science. This study would not have been possible without the dedicated efforts of University of Edinburg, The Centre for Inflammation Research laboratory scientists and the Generative AI Laboratory team. We would also like to thank Adiso Therapeutics for the provision of ADS032 and their team in collaboration: Christopher Murphy, PhD, Bharat Bixit, PhD, Jillian Chapas-Reed, BS, and Benjamin W. Miller, PharmD.

## Supplemental data

**Supp Table 1.**
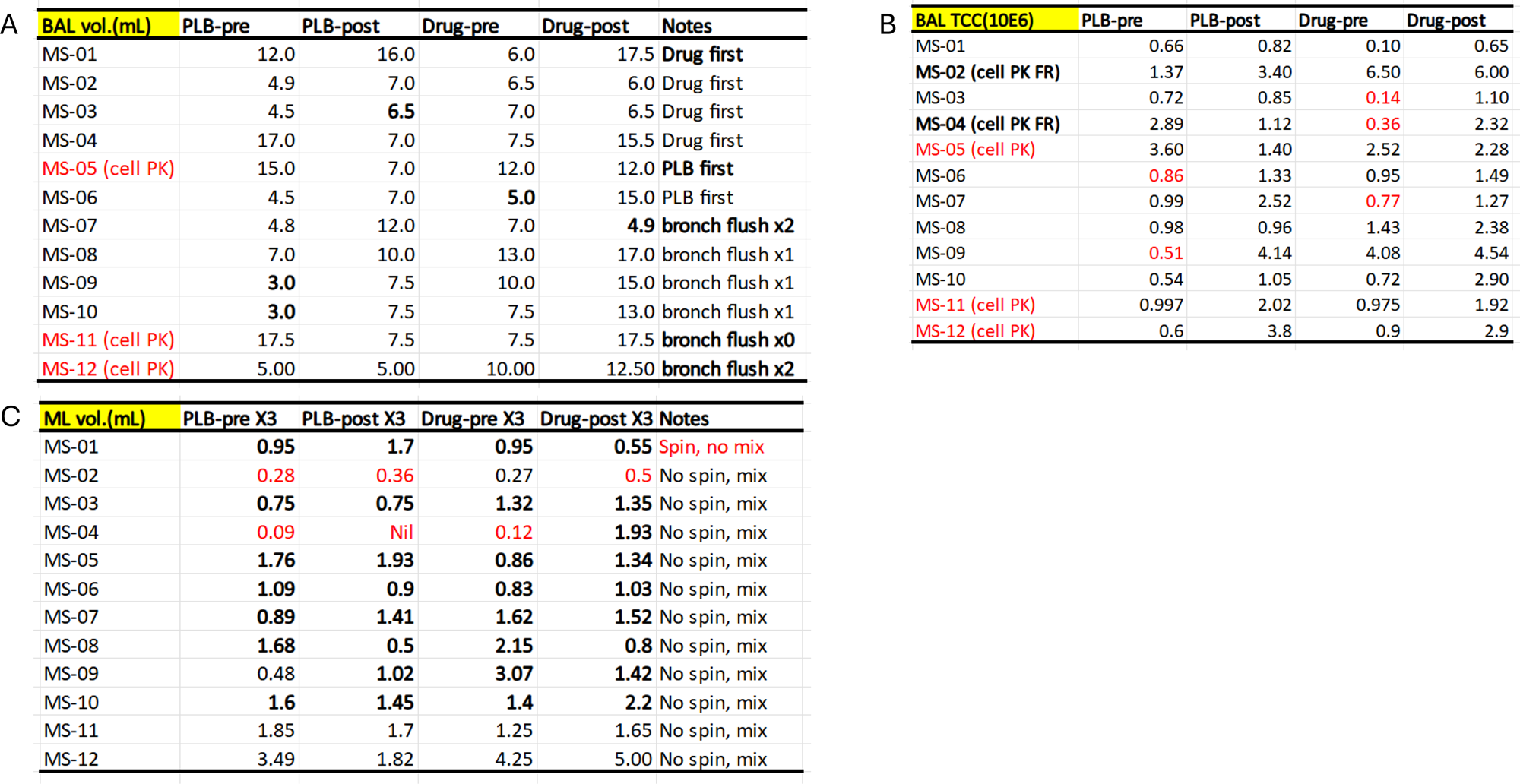
Tables displaying volumes of fluid and cell numbers obtained during Micro clinical trial. A: Bronchoalveolar lavage (BAL) volumes in microlitres (mL) for pre and post placebo and ADS032 lobes. B: Cell numbers (expressed in 10^6) obtained from BALs in pre and post placebo and ADS032 lobes. C: Microlavage volumes in microlitres (mL) for pre and post placebo and ADS032 lobes.

| BAL vol.(mL) | PLB-pre | PLB-post | Drug-pre | Drug-post | Notes |
| --- | --- | --- | --- | --- | --- |
| MS-01 | 12.0 | 16.0 | 6.0 | 17.5 | **Drug first** |
| MS-02 | 4.9 | 7.0 | 6.5 | 6.0 | Drug first |
| MS-03 | 4.5 | **6.5** | 7.0 | 6.5 | Drug first |
| MS-04 | 17.0 | 7.0 | 7.5 | 15.5 | Drug first |
| MS-05 (cell PK) | 15.0 | 7.0 | 12.0 | 12.0 | **PLB first** |
| MS-06 | 4.5 | 7.0 | **5.0** | 15.0 | PLB first |
| MS-07 | 4.8 | 12.0 | 7.0 | **4.9** | **bronch flush x2** |
| MS-08 | 7.0 | 10.0 | 13.0 | 17.0 | bronch flush x1 |
| MS-09 | **3.0** | 7.5 | 10.0 | 15.0 | bronch flush x1 |
| MS-10 | **3.0** | 7.5 | 7.5 | 13.0 | bronch flush x1 |
| MS-11 (cell PK) | 17.5 | 7.5 | 7.5 | 17.5 | **bronch flush x0** |
| MS-12 (cell PK) | 5.00 | 5.00 | 10.00 | 12.50 | **bronch flush x2** |

C
| ML vol.(mL) | PLB-pre X3 | PLB-post X3 | Drug-pre X3 | Drug-post X3 | Notes |
| --- | --- | --- | --- | --- | --- |
| MS-01 | **0.95** | **1.7** | **0.95** | **0.55** | **Spin, no mix** |
| MS-02 | **0.28** | **0.36** | 0.27 | **0.5** | No spin, mix |
| MS-03 | **0.75** | **0.75** | **1.32** | **1.35** | No spin, mix |
| MS-04 | **0.09** | **Nil** | **0.12** | **1.93** | No spin, mix |
| MS-05 | **1.76** | **1.93** | **0.86** | **1.34** | No spin, mix |
| MS-06 | **1.09** | **0.9** | **0.83** | **1.03** | No spin, mix |
| MS-07 | **0.89** | **1.41** | **1.62** | **1.52** | No spin, mix |
| MS-08 | **1.68** | **0.5** | **2.15** | **0.8** | No spin, mix |
| MS-09 | 0.48 | **1.02** | **3.07** | **1.42** | No spin, mix |
| MS-10 | **1.6** | **1.45** | **1.4** | **2.2** | No spin, mix |
| MS-11 | 1.85 | 1.7 | 1.25 | 1.65 | No spin, mix |
| MS-12 | 3.49 | 1.82 | 4.25 | 5.00 | No spin, mix |

- **Supp Table 1.** Tables displaying volumes of fluid and cell numbers obtained during Micro clinical trial.
| BAL TCC(10E6) | PLB-pre | PLB-post | Drug-pre | Drug-post |
| --- | --- | --- | --- | --- |
| MS-01 | 0.66 | 0.82 | 0.10 | 0.65 |
| MS-02 (cell PK FR) | 1.37 | 3.40 | 6.50 | 6.00 |
| MS-03 | 0.72 | 0.85 | **0.14** | 1.10 |
| MS-04 (cell PK FR) | 2.89 | 1.12 | **0.36** | 2.32 |
| MS-05 (cell PK) | 3.60 | 1.40 | 2.52 | 2.28 |
| MS-06 | **0.86** | 1.33 | 0.95 | 1.49 |
| MS-07 | 0.99 | 2.52 | **0.77** | 1.27 |
| MS-08 | 0.98 | 0.96 | 1.43 | 2.38 |
| MS-09 | **0.51** | 4.14 | 4.08 | 4.54 |
| MS-10 | 0.54 | 1.05 | 0.72 | 2.90 |
| MS-11 (cell PK) | 0.997 | 2.02 | 0.975 | 1.92 |
| MS-12 (cell PK) | 0.6 | 3.8 | 0.9 | 2.9 |

**Supp. Figure 1:**
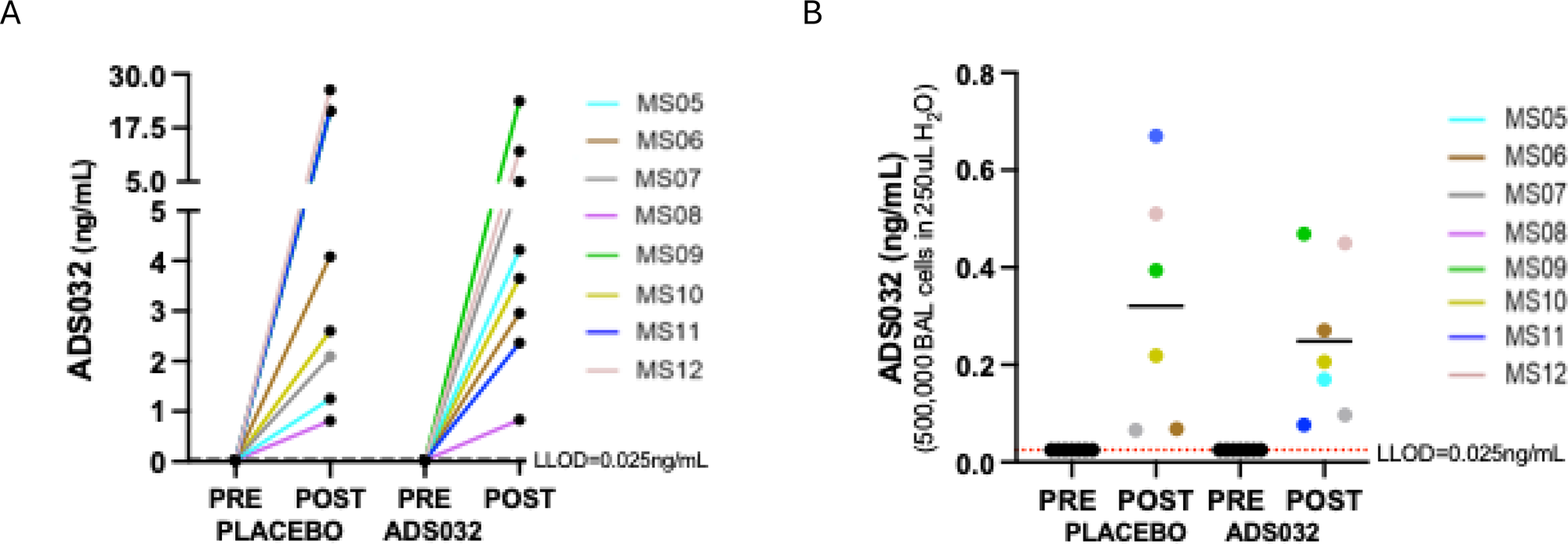
A&B: ADS032 concentration as measured by LC-MS. A: ADS032 concentration in bronchoalveolar lavage (BAL) fluid. All baseline pre placebo and pre ADS032 samples were below the LLOQ. No statistical difference between post placebo and post ADS032 BAL samples. Only participants MS08-MS12 included. B: ADS032 concentration in cells isolated from BALs. All pre placebo and pre ADS032 were below LLOQ. MS05 post placebo sample not available. No statistical difference between post placebo and post ADS032 BAL cell samples.

**Supp data Fig. 2a.**
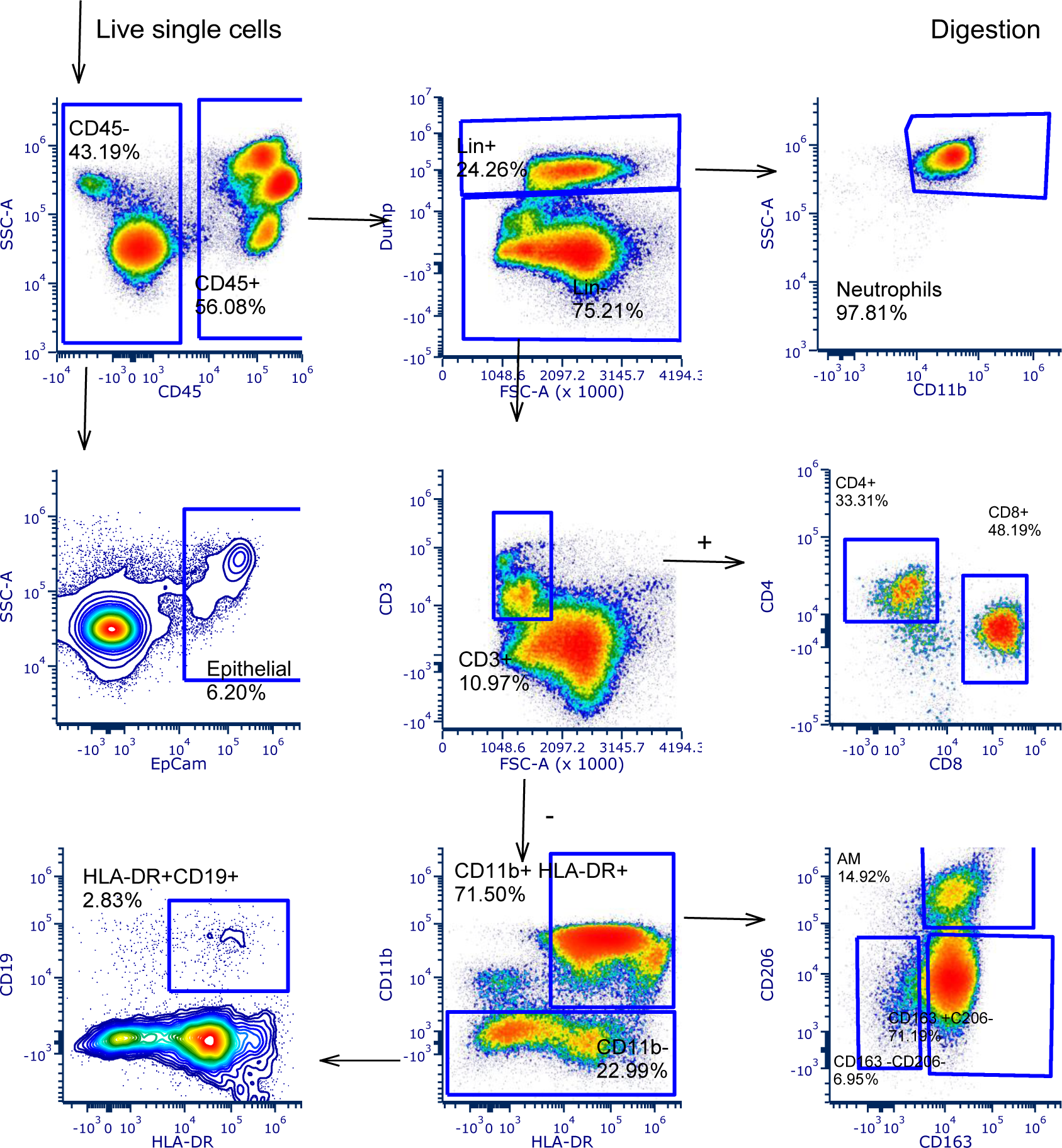
Flow cytometry gating strategy for alveolar macrophages, neutrophils, lymphocytes, and epithelial cells. Lung tissue digest strategy.

**Supp data Fig. 2b.**
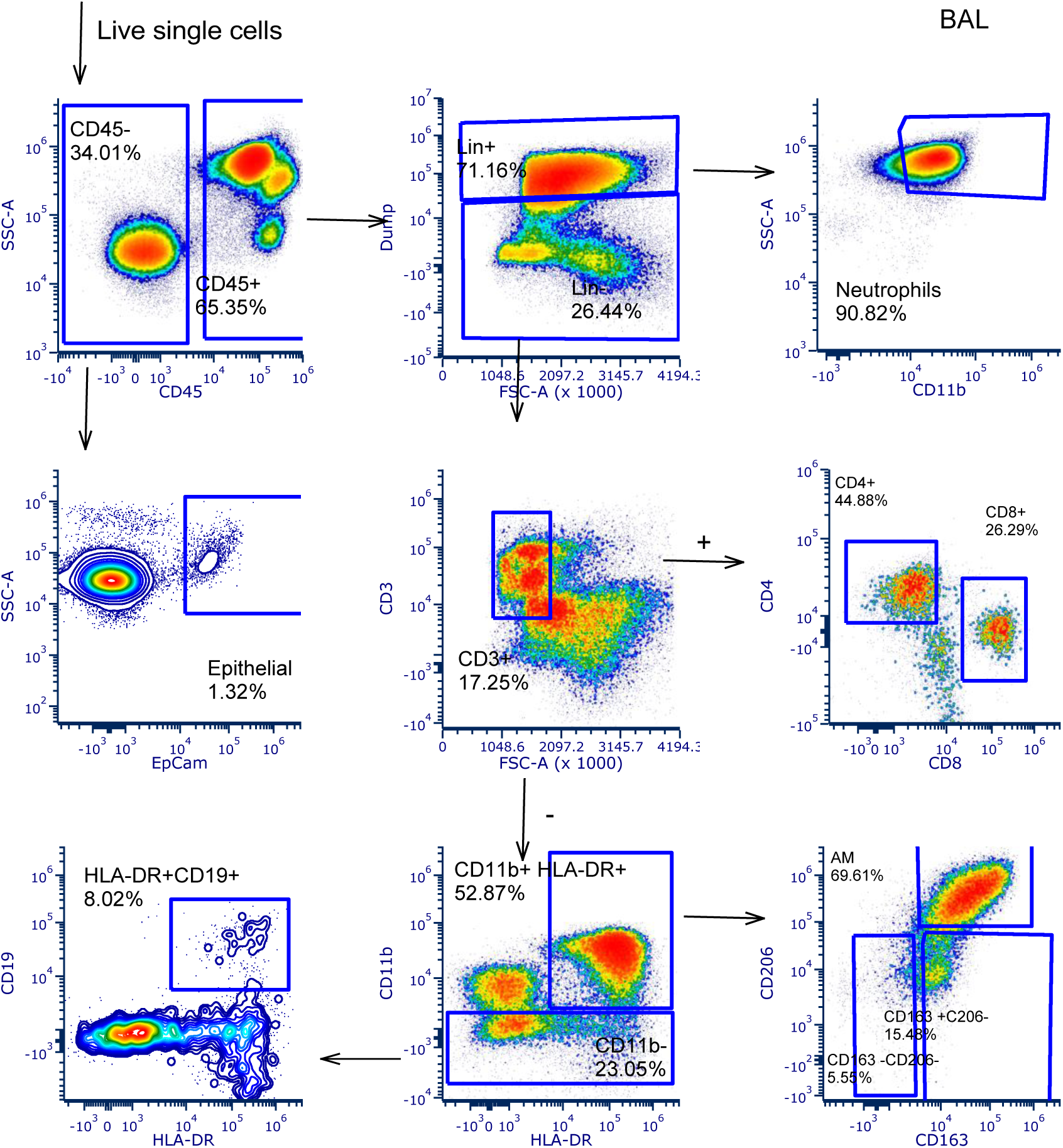
Flow cytometry gating strategy for alveolar macrophages, neutrophils, lymphocytes, and epithelial cells. Bronchoscopic lavage strategy.

**Supp Data Fig. 3.**
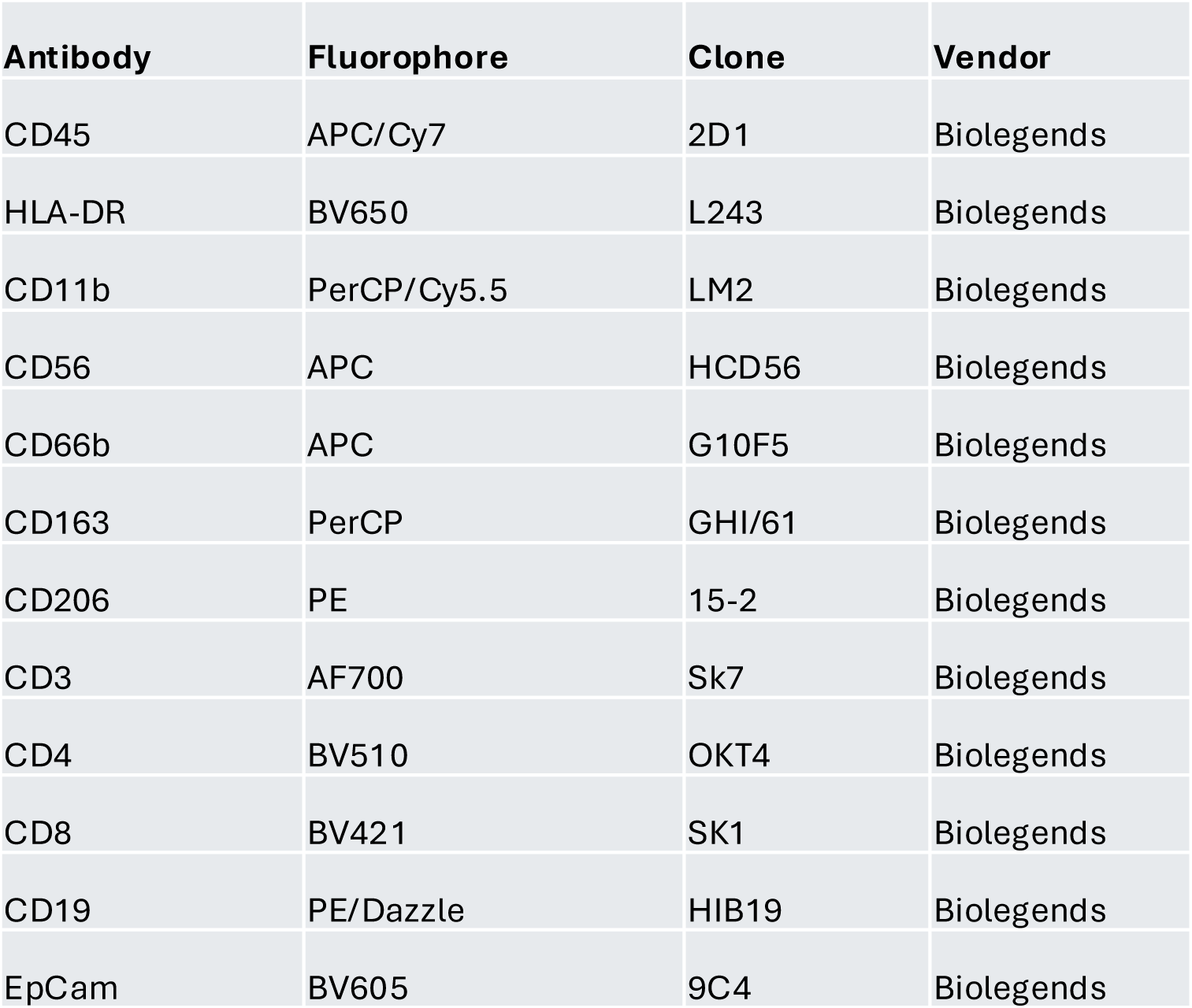
List of antibodies used in Flow cytometry analysis

**Supp Data Fig. 4.**
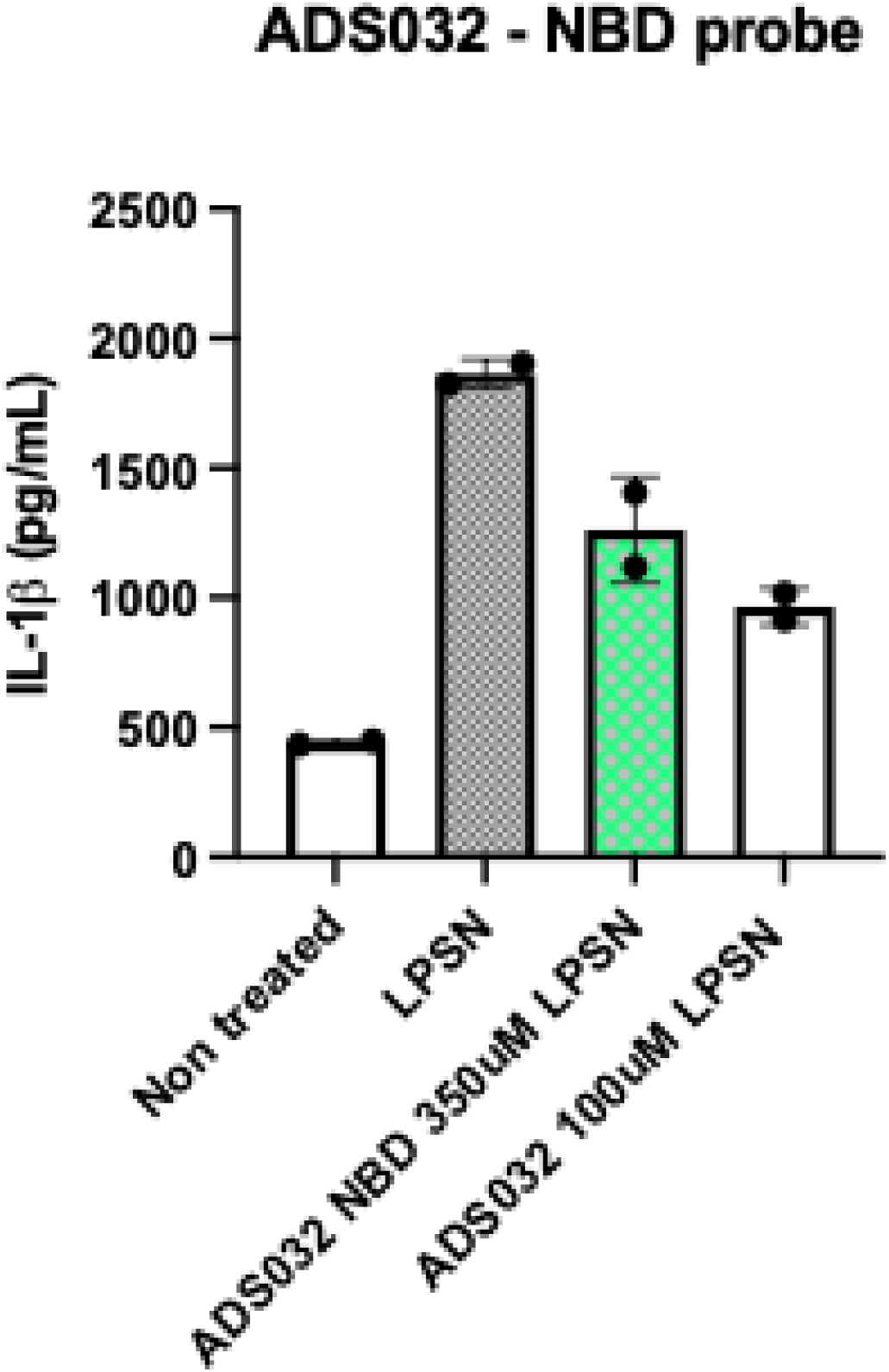
Comparative potency of ADS032 and ADS032–NBD in primary human alveolar macrophages. ADS032-NBD retains a degree of inhibitory ability. IL-1β release from AMs.

**Supp Data Fig. 5.**
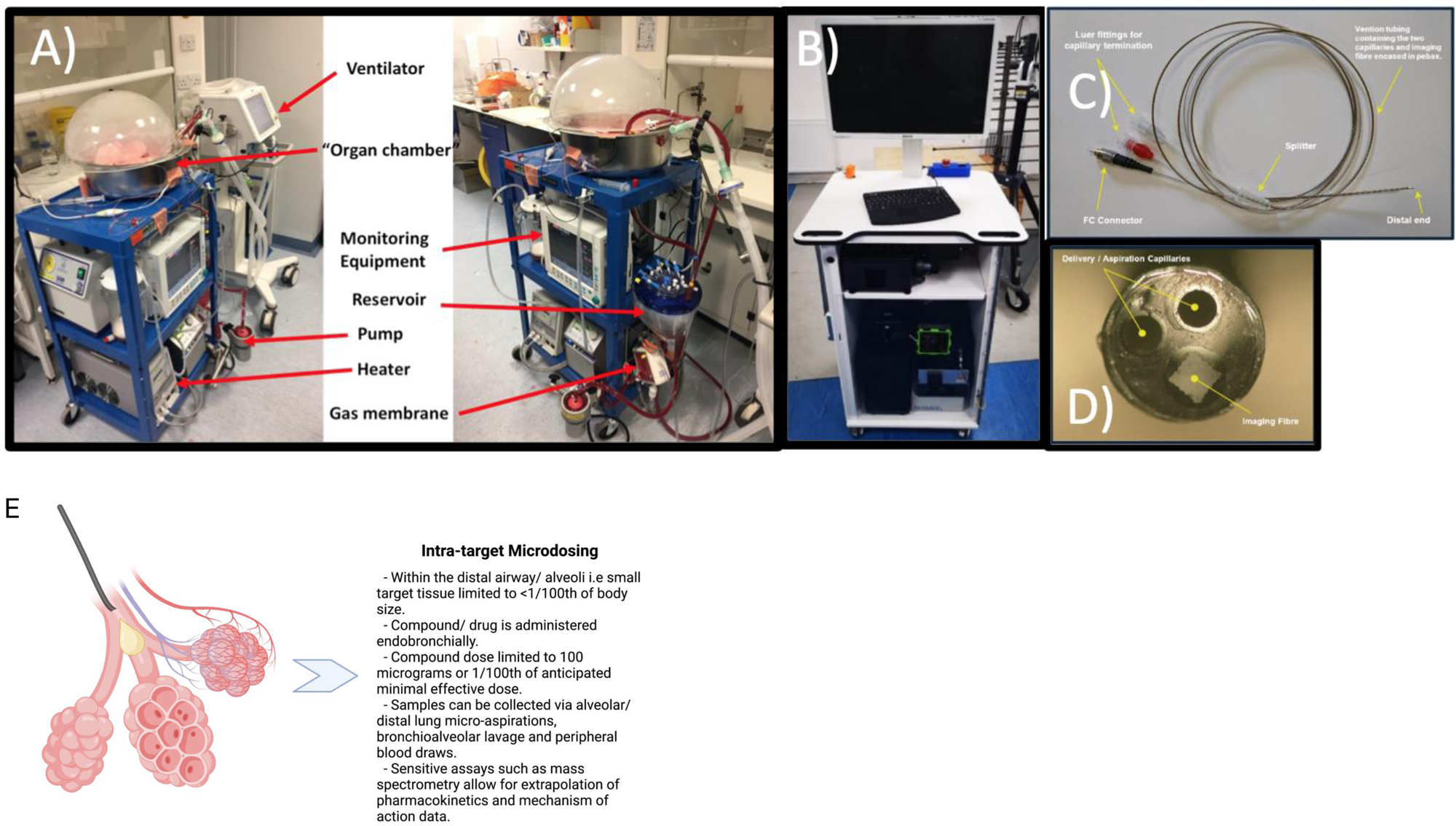
Ex vivo lung perfusion system setup and intrapulmonary delivery schematic. Ex Vivo Lung Perfusion, KronoScan Imaging Device & Panoptes Imaging Fibre and in situ detection of target drug engagment. A) Ex Vivo Lung Ventilation (EVLV). Human lungs were ventilated / perfused within the EVLV rig. B) KronoScan Imaging Device. C) Panoptes imaging fibre. Imaging fibre consists of 2 delivery/aspiration capillaries alongside imaging core containing over 8000 individual imaging fibres. Using a bronchoscope, the Panoptes imaging fibre can be navigated to the distal airways to deliver fluorescent imaging agents and visualise targets of interest. D) Magnified image of Panoptes imaging fibre. E) Schematic explaining the process of intra target microdosing within the distal lung.

