## Supplemental methods for "First-in-human intrapulmonary intratarget microdosing of a novel dual inflammasome inhibitor of NLRP 1/ NLRP 3 in ex vivo human lungs and patients with interstitial lung disease"

**Liquid Chromatography Mass spectrometry Materials and Methods**

**METHODS and Application –** Scott G Denham, Sofia Laforest, Stephen Kaluzny, Natalie ZM Homer

**Chemicals, reagents and consumables**

Water (LC-MS grade), acetonitrile (LC-MS grade), methanol (LC-MS grade) were from VWR, Lutterworth, UK. Formic acid (LC-MS grade) was from Fisher Scientific, Loughborough, UK).

Analytical standard for ADS032 (lot EW16223-242-P1) was supplied by Adiso as was certified reference material for isotopically labelled 2,2,4,4,6,6,21,21,21 -[^2^H]_9_ -progesterone (d9-P4) 100 µg/mL in acetonitrile. Powdered ADS032 was stored at room temperature while prepared solutions of ADS032 and d9-P4 were stored at -20^o^C.

ISOLUTE^®^, PLD+ 96-well plates (918-0050-P01) and 2 mL deep well collection plates (121-5203) were from Biotage (Uppsala, Sweden) and 2 mL deep well collection plates were from Waters (Wilmslow, UK; 121-5203). The BEH C18 50 x 2.1 mm, 1.7 µm liquid chromatography column was supplied by Waters (Wilmslow, UK; 186009455) and fitted with a Premier BEH VanGuard Fit Cartridges (5 x 2.1 mm, 1.7 µm; 186009459) Waters, UK.

**Stock solutions and calibration standards**

Standard solutions were prepared by dissolving a suitable amount of ADS032 powder in methanol (1 mg/mL). Further serial dilutions of the ADS032 were prepared in methanol from 50 µg/mL to 500 pg/mL. Isotopically labelled internal standard (d9-P4) was prepared by dilution to 25 ng/mL in methanol. Calibration standards were prepared freshly for each batch of samples analysed. A quality control (QC) pool was prepared and analysed with each batch to assess batch variability.

**Automated Sample Extraction of biological samples using a liquid handling robot**

Biological samples (plasma, urine, alveolar lavage (Micro lavage), bronchial alveolar lavage (BAL) fluid, and supernatant from cell lysates, as well as *ex vivo* lung perfusate) calibration standards (15 points, ranging from 0.025 – 10 ng/mL, quality control (QC) samples and blanks were dispensed manually as aliquots (100 µL) into individual wells of a 2mL 96 deep-well polypropylene plate (Waters, UK). The internal standard solution of D9-P4 in methanol was added (20 µL; 0.5 ng) to each well except for the double blanks. The plate was agitated on a plate shaker (2 mins) and transferred to a Biotage^®^ Extrahera^™^ automated sample processor (Biotage, Uppsala, Sweden) where the solution was transferred to a PLD+ plate by the robot and acetonitrile with 0.1% formic acid v/v (400 µL) was added to each well. Samples were incubated at room temperature (18‑22 ^°^C; 10 mins). Analytes were eluted from the PLD+ material into a deep-well collection plate by positive pressure. The eluate was reduced to dryness under a stream of heated oxygen free nitrogen (OFN, 40 °C) on an SPE Dry^TM^ Dual Sample Concentrator System (Biotage, Uppsala, Sweden). Once dry the extracts were dissolved in water/acetonitrile (70:30, 80 µL), the plate was sealed and shaken on a plate shaker (10 mins) before injecting (20 µL) directly from the 96-well plate for LC-MS/MS analysis.

**Chromatographic and Mass Spectrometric conditions for plasma steroid profiling**

LC-MS/MS analysis was performed using an Acquity I-Class UPLC (Waters, UK) fitted with a BEH C18 50 x 2.1 mm, 1.7 µm liquid chromatography column supplied by Waters (Wilmslow, UK; 186009455) interfaced to a QTRAP 6500+ (AB Sciex, Warrington, UK) mass spectrometer. Instrument control and data acquisition were achieved using Sciex Analyst^®^ 1.7.3 Software. Data were integrated using software package Sciex MultiQuant 3.0.3.

Chromatographic separation was achieved on the BEH C18 50 x 2.1 mm, 1.7 µm liquid chromatography column using mobile phase A - water (0.1% formic acid v/v) and mobile phase B - acetonitrile (0.1% formic acid v/v) at a flow rate of 0.4 mL/min with a 6 minute total gradient starting at 30% B for 0.5 mins, rising to 100% B up to 3.5 mins, held for 0.9 mins, returned to 30% B over 0.1 min and re-equlibrated for 1.5 mins at a column temperature of 50^o^C.

The mass spectrometer was operated in positive ion in electrospray ionisation (ESI) mode using a TurboIonSpray source and data were collected in unit resolution (0.7 *m/z* full width at half maximum). The ESI source was operated at 600^o^C with a voltage of +5 kV, a Curtain Gas of 40 psi, nitrogen nebuliser ion source gas 1 and heater ion source gas 2 of 40 psi and 60 psi, respectively. Compound specific parameters were optimised for multiple reaction monitoring (MRM) transitions by infusing 100 ng/mL of ADS032 and D9-P4 standard solution into the source. The curtain, source, exhaust and CAD gas was delivered using an MS Table 1N Nitrogen and dry air generator Table (Peak Scientific, Scotland, UK). Two transitions were selected for each steroid – the quantitative and qualitative ion - and their optimised declustering potential, collision exit potential and collision energy are listed in Supplementary Table 1.

| **Supplementary Table 1 – Retention time and Mass Spectrometry parameters for positive ion MRM on an Acquity I-Class UPLC, C18 BEH column, coupled to QTrap 6500+ mass spectrometer** | | | | | | | | |
| --- | --- | --- | --- | --- | --- | --- | --- | --- |
| **Q1 Mass** | **Q3 Mass** | **Retention time (min)** | **Transition ID** | **DP (volts)** | **CE (volts)** | **CXP (volts)** | **Polarity** | **Internal Standard** |
| 404.1 | 302.9 | 2.64 | ADS032 1 | 51 | 23 | 26 | + | D9P4 |
| 404.1 | 238.9 | 2.64 | ADS032 2 | 51 | 31 | 28 | + | D9P4 |
| 324.1 | 100.0 | 2.45 | D9-Progesterone (D9-P4) | 96 | 23 | 10 | + |  |

**Chromatographic Data Handling**

The data was analysed on a quantitative chromatographic software package called MultiQuant v3.0.3 (Sciex, UK). Quantification of ADSO32 was performed following integration of the chromatographic peaks using linear regression of the peak area ratio (PAR) of ADS032 to d9P4 in the calibration standards, QCs and unknowns. The PAR of the analyte (ADS032) and the internal standard (d9P4) were plotted against concentration of ADS032 and the amount of ADS032 was calculated in unknowns and QCs from the linear regression of the equation.

The ratio of the peak area of the quantitative ion (m/z 404.1 🡪 302.9) at the retention time 2.64 mins to the qualitative ion (m/z 404.1 🡪 238.9) at the same retention time was also calculate, to ensure the analyte does not have interference in the signal. Quantitative/qualitative ion ratio was considered acceptable based on the ion ratio of the calibration standards. Any peak in a biological sample that had an ion ratio that exceeded 20 % was excluded from the calculation of ADS32 quantity and the result was not compiled into the final data set. The data was extracted into excel for final calculation of concentration in the sample.

### Limits of Quantitation, precision and recovery of the steroid profiling method for plasma

Standard curves were assessed with acceptable linearity and a mean r value of 0.99. Best fit was achieved using weighting of 1/x. Recovery and matrix effects were assessed in plasma. Assay precision was evaluated by comparing replicates of quality control samples between day (inter-assay variation) (n=3).

| **Supplementary Table 2 - Validation criteria showing linear range, recovery and inter-assay precision for steroid profiling of chicken plasma with analysis on an Acquity IClass UPLC fitted with a C18 BEH column, coupled to a QTrap 6500+ mass spectrometer** | | | | | | |
| --- | --- | --- | --- | --- | --- | --- |
| **Analyte** |  | **Linear Range (ng/mL)** | **Linearity (R)** | **Matrix effects (%)** | **Recovery from plasma (%)** | **Inter-day**  **Precision** |
| ADS032 | LLOQ | 0.0025-50 | 0.999 | 45% | 68 | 4.9 |
